# Predictors of functional outcome following acute ischemic stroke secondary to M1 MCA/ICA occlusion in the extended time window

**DOI:** 10.1101/2024.11.05.24316799

**Authors:** John Constantakis, Quinn Steiner, Thomas Reher, Timothy Choi, Fauzia Holangel, Qianqian Zhao, Nicole Bennett, Veena A. Nair, Eric Adelman, Vivek Prabhakaran, Beverly Aagard-Kienitz, Bolanle Famakin

## Abstract

**Introduction:** A validated clinical decision tool predictive of favorable functional outcomes following endovascular thrombectomy (EVT) in acute ischemic stroke (AIS) remains elusive. We performed a retrospective case series of patients at our regional Comprehensive Stroke Center, over a 4-year period, who have undergone EVT to elucidate patient characteristics and factors associated with a favorable functional outcome after EVT.

**Methods:** We reviewed all cases of EVT at our institution from 2/2018 - 2/2022 in the extended time window from 6 – 24 hours. Demographic, clinical, imaging, and procedure co-variates were included. A favorable clinical outcome was defined as a modified Rankin scale or 0-2. We included patients with M1 or ICA occlusion treated with EVT within 6-24 hours after symptom onset. We used a univariate and multivariate logistic regression analysis to identify patient factors associated with a favorable clinical outcome at 90 days.

**Results:** Our analysis demonstrates that higher recanalization score based on the mTICI scale (2B-3) was a strong indicator of favorable outcome per both our multivariate and univariate analysis (OR 4.11; CI 1.10 – 15.31; p 0.035). Our data also showed signal that the younger age (*p* 0.013), lower baseline NIHSS (*p* 0.043), shorter hospital length of stay (LOS) (*p* 0.030), and absence of pre-existing hypertension (*p* 0.026) may also be a predictor of favorable outcome per our univariate analysis.

**Conclusion:** Patients without pre-existing hypertension had more favorable outcomes following EVT in the expanded time window. This is consistent with other multicenter data in the expanded time window that demonstrates greater odds of a poor outcome with elevated pre-, peri-, and post-endovascular treatment blood pressure. Our data also demonstrates mTICI score is a strong predictor of favorable outcome even when controlled for other variables. Other factors that may indicate a favorable outcome include younger age, lower baseline NIHSS, and shorter hospital LOS.

## Introduction

Ischemic stroke incidence, prevalence, and mortality are increasing annually with this trend expected to continue^1^. Endovascular intervention for stroke has emerged as an effective treatment strategy in patients with an acute ischemic stroke secondary to large vessel occlusion (LVO)^2^. Initially, significant clinical benefit was seen when endovascular intervention was performed within 6 hours after the onset of stroke symptoms^3,4^. However, the recent DEFUSE-3 and DAWN trials expanded this time window out to 6-24 hours^5,6^.

Although the guidelines for the acute treatment of stroke have evolved to include this expanded time window from 6-24 hours; even according to our study, there are large variations in patient outcomes. Within this expanded time window, 40-50% of patients have poor neurological outcomes per modified Rankin scores (mRS) (defined as mRS 3-6), with mortality rates around 15%^5-8^. A variety of factors have been identified as correlating with worse clinical outcomes following stroke^9,10^; however, a clinically usable model for outcome prognostication has remained elusive.

We sought to examine our single center experience at a major, regional, comprehensive stroke center with endovascular intervention for acute ischemic stroke in patients with M1 MCA and ICA occlusions. Patients with these types of LVOs typically have a worse prognosis^5,6^. These LVOs constitute many patients that undergo endovascular intervention^5,6^. Consequently, understanding factors that impact outcomes may greatly improve quality of care.

The purpose of our investigation is to identify potential clinical and imaging biomarkers predictive of a favorable functional outcome following acute stroke secondary to M1 and ICA occlusion during the extended time window. This would ideally be used to predict pre-procedurally which patients would show the greatest functional outcome response to thrombectomy.

## Methods

### Patient Selection

After approval from our institutional review board, we retrospectively reviewed all patients admitted or transferred to our hospital who underwent endovascular stroke treatment for arterial ischemic stroke from 6 – 24 hours after symptom onset between 2/2018 - 2/2022. The decision to perform the endovascular procedure was made on an individual basis with consensus between the neuro-interventionalist and neurology team. We included patients older than 18 years who underwent endovascular intervention for occlusion of M1 vessels as well as either intracranial or cervical ICA vessels and had baseline imaging performed at University of Wisconsin University Hospital, had a time from symptoms onset of greater than 6 but less than 24 hours. All procedures were performed using standard of care guidelines^5,6^. We collected patient baseline clinical and radiological characteristics, procedure details, and outcomes such as the degree of successful recanalization according to the modified Thrombolysis In Cerebral Infarction [mTICI] scale (grade of 0-3, with higher scores indicating greater recanalization). Specifically, a grade is assigned at the end of the procedure-[mTICI 2b and less indicating lesser recanalization, and rates of complete/near-complete recanalization defined as [mTICI 3, mTICI 2c]. We defined a favorable outcome as an mRS score of 0-2 at 90-days post presentation^11^. We also compared whether each patient taken for thrombectomy met criteria as outlined by the DAWN and DEFUSE-3 trials in terms of clinical or radiographic mismatch.

### Statistical Methodology

#### Outcome Analyses

To assess the predictors of a patient encountering an unfavorable outcome, we conducted a univariate and multivariate logistic regression model and reported odds ratios and 95% confidence intervals. Significant predictors from the univariate model were adjusted for in the multivariate analyses. We graphically examined differences in the outcome using coefficient plots with extended confidence widths. We checked for multicollinear variables using a variance inflation factor (VIF). Any variable with a VIF values ≥10 was excluded from the multivariate analyses. We corrected for missing data using pairwise deletion for categorical data or multiple imputation methods for continuous variables. Variables with greater than 50% missing data were removed from the analysis altogether. All p-values less than 0.05 were considered significant. All statistical analyses were completed with the use of SAS Version 9.4 (Cary, NC) and STATA version 17.

#### Descriptive Analyses

We summarized demographic and outcome measures using raw counts and frequencies for all categorical, binary, and ordinal variables. Comparisons between favorable outcome groups were made using chi-square tests or Fisher’s exact tests for cells less than 5. Continuous variables were expressed as mean plus standard deviation (SD) or median plus interquartile range (IQR) for non-normal data. Continuous variable comparisons were made using a student t-test or the non-parametric Wilcoxon rank sum test for non-normal distributions.

## Results

A total of 679 patients underwent thrombectomy at our comprehensive stroke center over a 4-year period. Of these, 121 (18%) met inclusion criteria (CT or MR perfusion on arrival, carotid or M1 occlusion) (Figure 1), and satisfactory data complements were found in 54 of these 121 (45%)-meaning the variable contained >50% of the investigated data fields.

**Figure 1:**
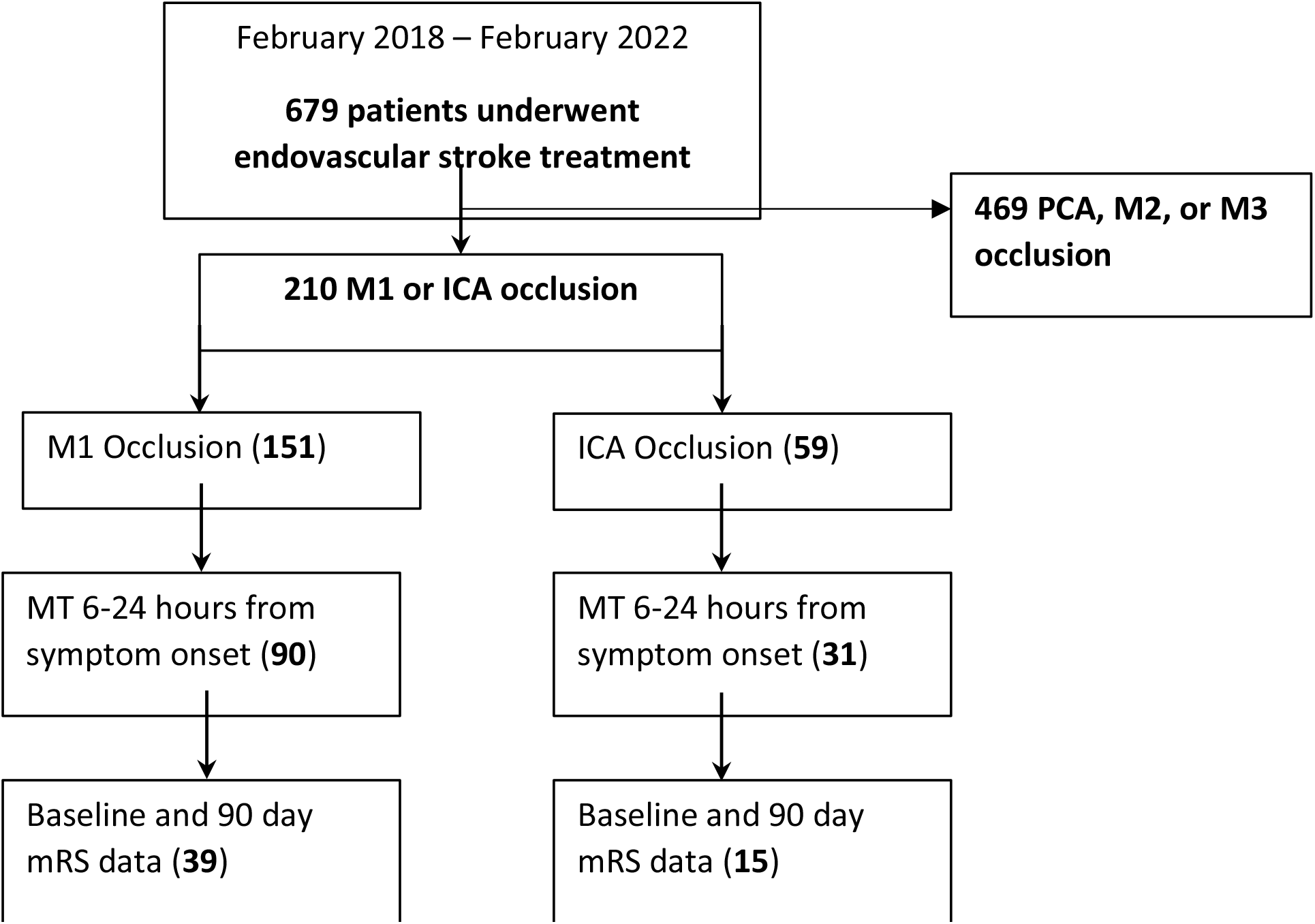
Patient Flow Chart.

### Patient Cohort Characteristics and Outcomes

Baseline characteristics are outlined in Table 1. The groups did not differ significantly in occlusion site, sex, or smoking history. The groups also did not differ significantly in whether they had a diagnosis of diabetes, atrial fibrillation, decreased cardiac ejection fraction (EF), or cervical carotid involvement. Favorable clinical outcomes were observed in 55 (44%) patients, 15 (18%) of which had ICA occlusions and 45 (82%) of which had M1 occlusions. Patients with favorable outcomes had a younger age, lower baseline NIHSS, a shorter length of stay, and higher TICI score.

**Table 1:**
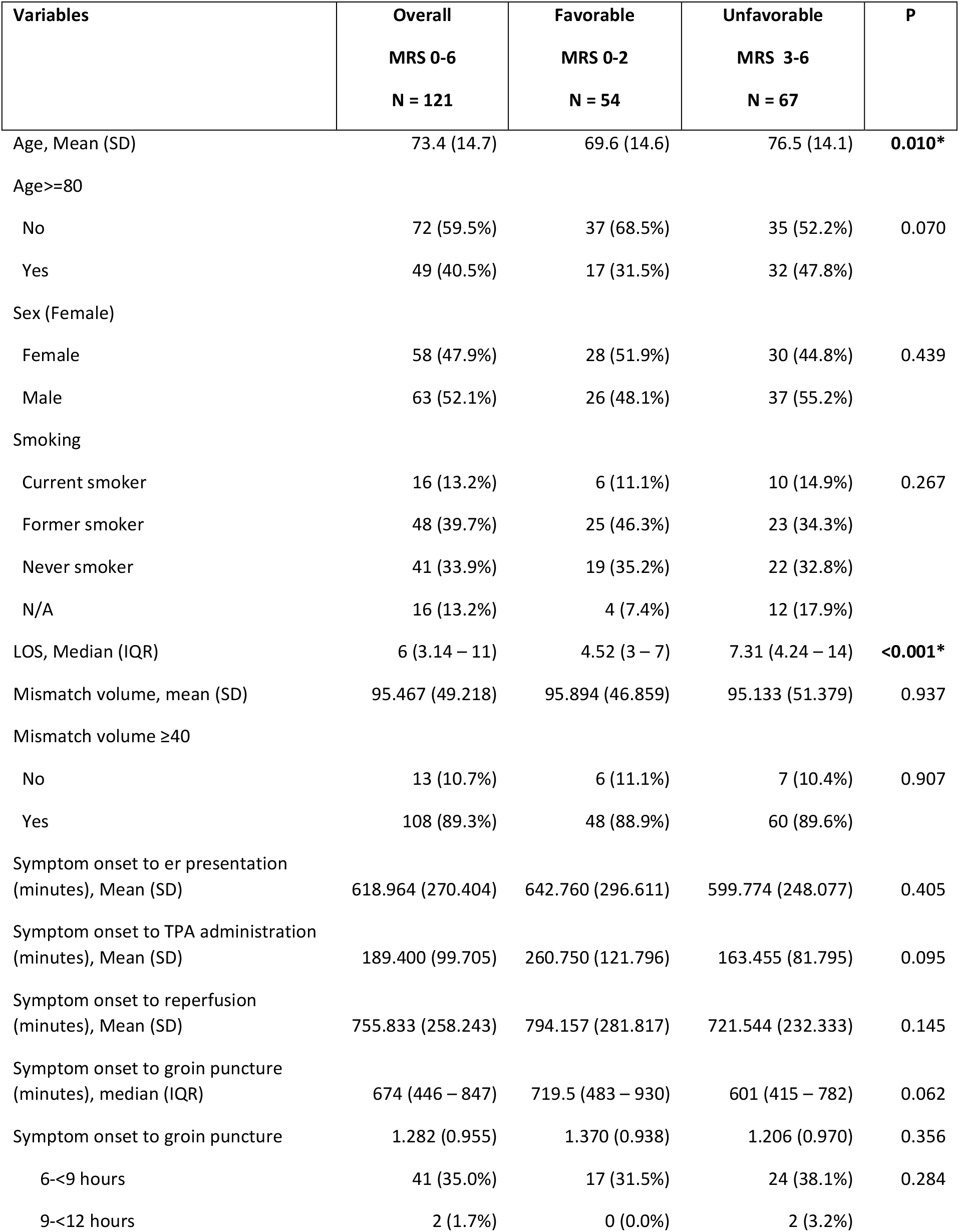
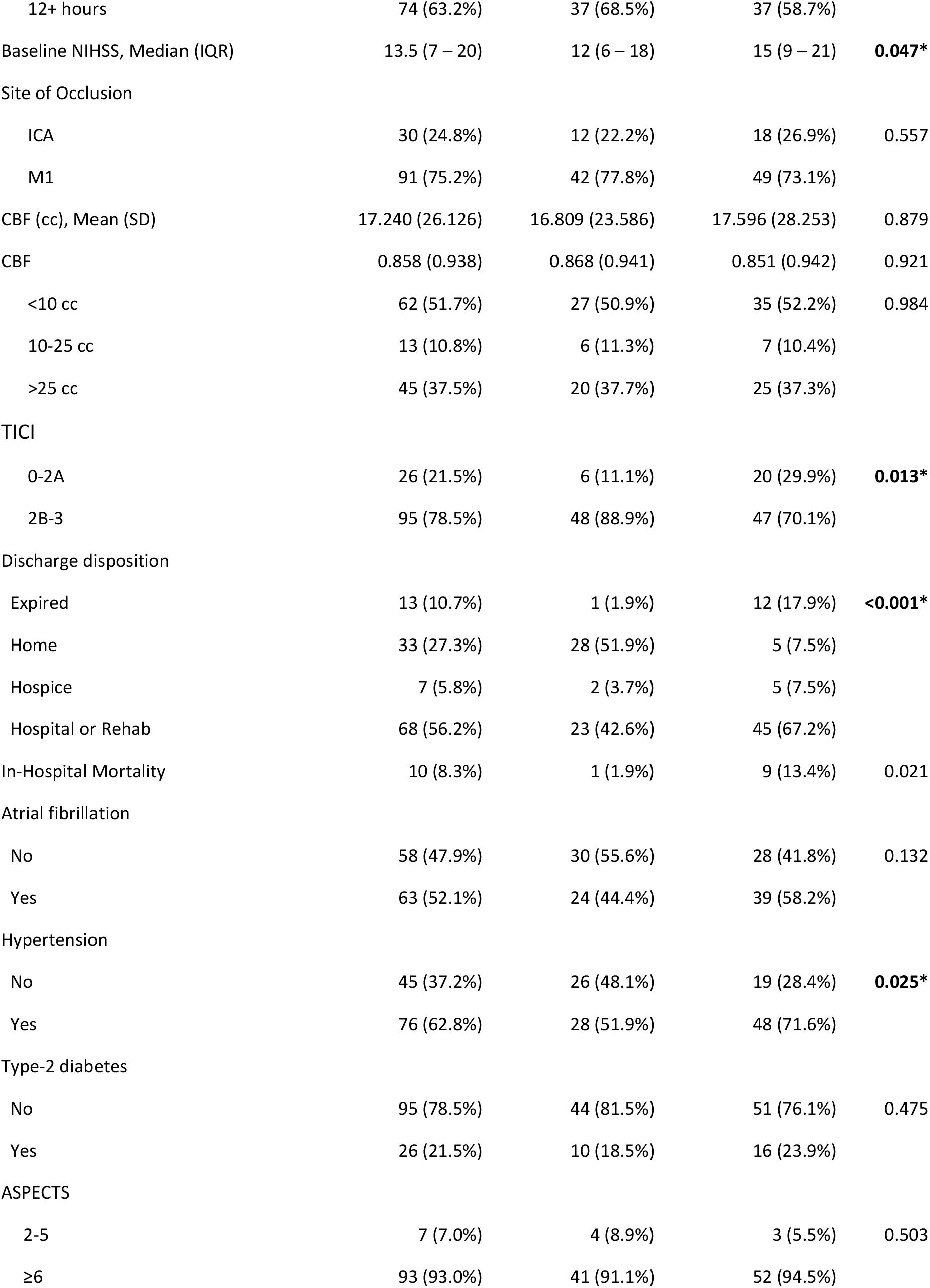
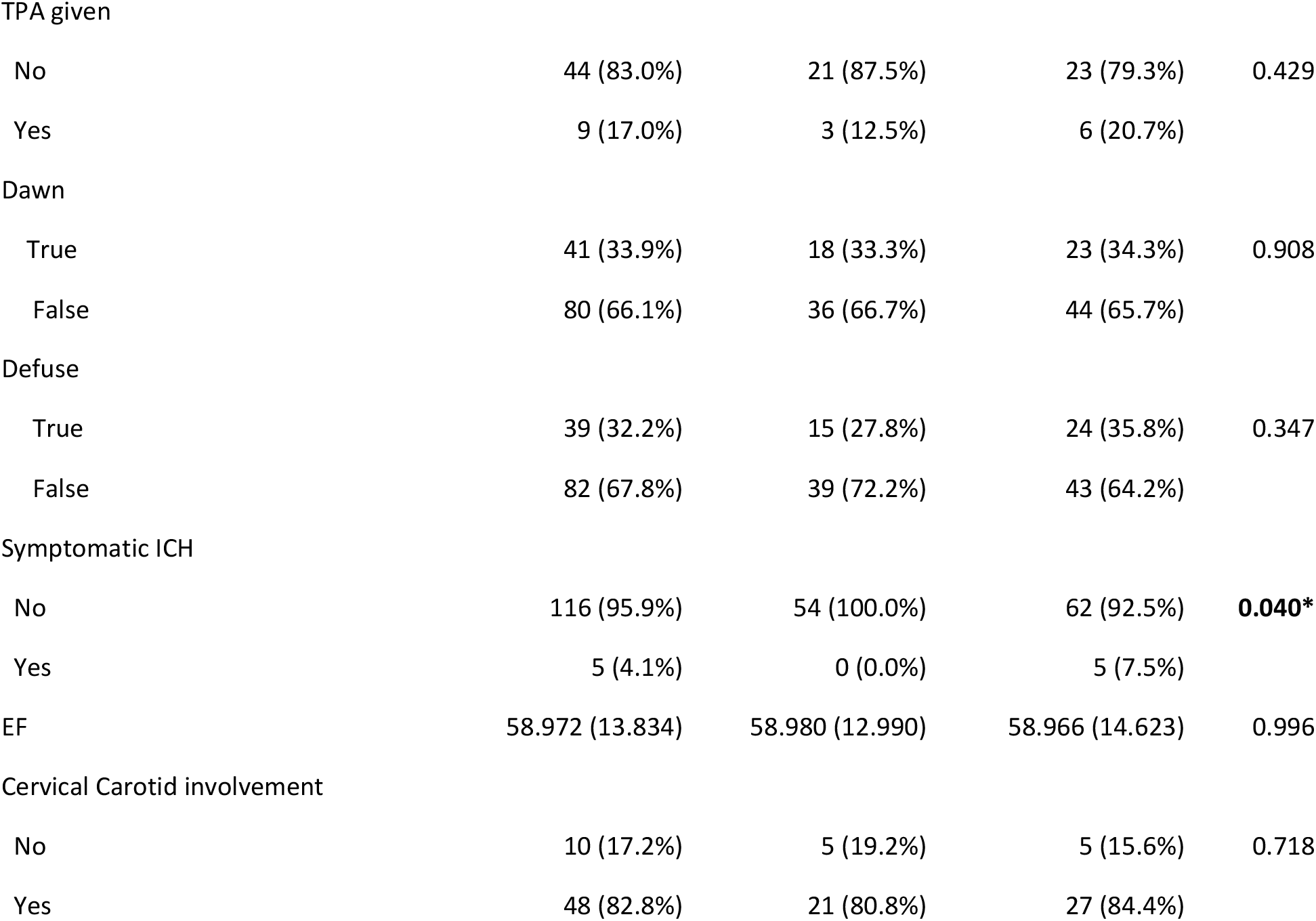
Baseline characteristics and medical history of cohort. This table presents baseline characteristics and medical history of a cohort, categorized by favorable (MRS 0-2) and unfavorable (MRS 3-6) outcomes, with an overall grouping of MRS 0-6.

| Variables | Overall<br>MRS 0-6<br>N = 121 | Favorable<br>MRS 0-2<br>N = 54 | Unfavorable<br>MRS 3-6<br>N = 67 | P |
| --- | --- | --- | --- | --- |
| Age, Mean (SD) | 73.4 (14.7) | 69.6 (14.6) | 76.5 (14.1) | **0.010\*** |
| Age≥80 |  |  |  |  |
| No | 72 (59.5%) | 37 (68.5%) | 35 (52.2%) | 0.070 |
| Yes | 49 (40.5%) | 17 (31.5%) | 32 (47.8%) |  |
| Sex (Female) |  |  |  |  |
| Female | 58 (47.9%) | 28 (51.9%) | 30 (44.8%) | 0.439 |
| Male | 63 (52.1%) | 26 (48.1%) | 37 (55.2%) |  |
| Smoking |  |  |  |  |
| Current smoker | 16 (13.2%) | 6 (11.1%) | 10 (14.9%) | 0.267 |
| Former smoker | 48 (39.7%) | 25 (46.3%) | 23 (34.3%) |  |
| Never smoker | 41 (33.9%) | 19 (35.2%) | 22 (32.8%) |  |
| N/A | 16 (13.2%) | 4 (7.4%) | 12 (17.9%) |  |
| LOS, Median (IQR) | 6 (3.14 – 11) | 4.52 (3 – 7) | 7.31 (4.24 – 14) | **<0.001\*** |
| Mismatch volume, mean (SD) | 95.467 (49.218) | 95.894 (46.859) | 95.133 (51.379) | 0.937 |
| Mismatch volume ≥40 |  |  |  |  |
| No | 13 (10.7%) | 6 (11.1%) | 7 (10.4%) | 0.907 |
| Yes | 108 (89.3%) | 48 (88.9%) | 60 (89.6%) |  |
| Symptom onset to er presentation<br>(minutes), Mean (SD) | 618.964 (270.404) | 642.760 (296.611) | 599.774 (248.077) | 0.405 |
| Symptom onset to TPA administration<br>(minutes), Mean (SD) | 189.400 (99.705) | 260.750 (121.796) | 163.455 (81.795) | 0.095 |
| Symptom onset to reperfusion<br>(minutes), Mean (SD) | 755.833 (258.243) | 794.157 (281.817) | 721.544 (232.333) | 0.145 |
| Symptom onset to groin puncture<br>(minutes), median (IQR) | 674 (446 – 847) | 719.5 (483 – 930) | 601 (415 – 782) | 0.062 |
| Symptom onset to groin puncture | 1.282 (0.955) | 1.370 (0.938) | 1.206 (0.970) | 0.356 |
| 6-<9 hours | 41 (35.0%) | 17 (31.5%) | 24 (38.1%) | 0.284 |
| 9-<12 hours | 2 (1.7%) | 0 (0.0%) | 2 (3.2%) |  |
| 12+ hours | 74 (63.2%) | 37 (68.5%) | 37 (58.7%) |  |
| Baseline NIHSS, Median (IQR) | 13.5 (7 – 20) | 12 (6 – 18) | 15 (9 – 21) | **0.047\*** |
| Site of Occlusion |  |  |  |  |
| ICA | 30 (24.8%) | 12 (22.2%) | 18 (26.9%) | 0.557 |
| M1 | 91 (75.2%) | 42 (77.8%) | 49 (73.1%) |  |
| CBF (cc), Mean (SD) | 17.240 (26.126) | 16.809 (23.586) | 17.596 (28.253) | 0.879 |
| CBF | 0.858 (0.938) | 0.868 (0.941) | 0.851 (0.942) | 0.921 |
| <10 cc | 62 (51.7%) | 27 (50.9%) | 35 (52.2%) | 0.984 |
| 10-25 cc | 13 (10.8%) | 6 (11.3%) | 7 (10.4%) |  |
| >25 cc | 45 (37.5%) | 20 (37.7%) | 25 (37.3%) |  |
| TICI |  |  |  |  |
| 0-2A | 26 (21.5%) | 6 (11.1%) | 20 (29.9%) | **0.013\*** |
| 2B-3 | 95 (78.5%) | 48 (88.9%) | 47 (70.1%) |  |
| Discharge disposition |  |  |  |  |
| Expired | 13 (10.7%) | 1 (1.9%) | 12 (17.9%) | **<0.001\*** |
| Home | 33 (27.3%) | 28 (51.9%) | 5 (7.5%) |  |
| Hospice | 7 (5.8%) | 2 (3.7%) | 5 (7.5%) |  |
| Hospital or Rehab | 68 (56.2%) | 23 (42.6%) | 45 (67.2%) |  |
| In-Hospital Mortality | 10 (8.3%) | 1 (1.9%) | 9 (13.4%) | 0.021 |
| Atrial fibrillation |  |  |  |  |
| No | 58 (47.9%) | 30 (55.6%) | 28 (41.8%) | 0.132 |
| Yes | 63 (52.1%) | 24 (44.4%) | 39 (58.2%) |  |
| Hypertension |  |  |  |  |
| No | 45 (37.2%) | 26 (48.1%) | 19 (28.4%) | **0.025\*** |
| Yes | 76 (62.8%) | 28 (51.9%) | 48 (71.6%) |  |
| Type-2 diabetes |  |  |  |  |
| No | 95 (78.5%) | 44 (81.5%) | 51 (76.1%) | 0.475 |
| Yes | 26 (21.5%) | 10 (18.5%) | 16 (23.9%) |  |
| ASPECTS |  |  |  |  |
| 2-5 | 7 (7.0%) | 4 (8.9%) | 3 (5.5%) | 0.503 |
| ≥6 | 93 (93.0%) | 41 (91.1%) | 52 (94.5%) |  |
| TPA given |  |  |  |  |
| No | 44 (83.0%) | 21 (87.5%) | 23 (79.3%) | 0.429 |
| Yes | 9 (17.0%) | 3 (12.5%) | 6 (20.7%) |  |
| Dawn |  |  |  |  |
| True | 41 (33.9%) | 18 (33.3%) | 23 (34.3%) | 0.908 |
| False | 80 (66.1%) | 36 (66.7%) | 44 (65.7%) |  |
| Defuse |  |  |  |  |
| True | 39 (32.2%) | 15 (27.8%) | 24 (35.8%) | 0.347 |
| False | 82 (67.8%) | 39 (72.2%) | 43 (64.2%) |  |
| Symptomatic ICH |  |  |  |  |
| No | 116 (95.9%) | 54 (100.0%) | 62 (92.5%) | **0.040\*** |
| Yes | 5 (4.1%) | 0 (0.0%) | 5 (7.5%) |  |
| EF | 58.972 (13.834) | 58.980 (12.990) | 58.966 (14.623) | 0.996 |
| Cervical Carotid involvement |  |  |  |  |
| No | 10 (17.2%) | 5 (19.2%) | 5 (15.6%) | 0.718 |
| Yes | 48 (82.8%) | 21 (80.8%) | 27 (84.4%) |  |

### Predictors of Favorable Outcomes

Based on the logistic regression analysis, several factors were examined to determine their association with favorable outcomes in the studied cohort. Age, Length of stay (LOS), and NIHSS were associated with favorable outcome in univariate analyses but were not significant predictors in the multivariable model.

Ejection fraction (EF) did not demonstrate a significant association with favorable outcomes in either univariate or multivariate analyses, suggesting that it may not be a strong predictor of favorable outcomes in this cohort. In summary, timely reperfusion therapy and discharge to home emerged as strong predictors of favorable outcomes, underscoring the importance of these interventions in improving outcomes for stroke patients. Although hypertension initially showed a significant association with favorable outcomes in the univariate analysis, this association became non-significant after adjusting for other variables, indicating that hypertension alone may not independently predict favorable outcomes. Other factors, such as age, LOS, stroke severity, hypertension, and EF, may not independently predict favorable outcomes once other variables are considered (Table 2).

**Table 2:** Predictors of favorable outcome (MRS 0 - 2) Logistic regression table presents predictors of a favorable outcome (MRS 0-2) after analyzing associated variables in both a univariate and multivariate analysis.

|  | Univariate |  |  | Multivariate |  |  |
| --- | --- | --- | --- | --- | --- | --- |
| Variables | OR | 95% CI | P | OR | 95% CI | P |
| Age, (years) | 0.97 | 0.94 – 0.99 | **0.013\*** | 0.98 | 0.95 – 1.02 | 0.274 |
| Sex (Female) | 1.33 | 0.65 – 2.72 | 0.439 |  |  |  |
| Smoking |  |  |  |  |  |  |
| Never smoker | Ref | Ref | Ref |  |  |  |
| Current smoker | 0.69 | 0.21 – 2.27 | 0.546 |  |  |  |
| Former smoker | 1.26 | 0.54 – 2.90 | 0.589 |  |  |  |
| LOS | 0.95 | 0.90 – 0.99 | **0.030\*** | 0.96 | 0.91 – 1.01 | 0.091 |
| Mismatch volume | 1.00 | 0.99 – 1.01 | 0.936 |  |  |  |
| Symptom onset to er presentation (minutes) | 1.00 | 0.99 – 1.00 | 0.402 |  |  |  |
| Symptom onset to TPA administration (minutes) | 1.01 | 0.99 – 1.03 | 0.151 |  |  |  |
| Symptom onset to reperfusion (minutes) | 1.00 | 0.99 - 1.00 | 0.146 |  |  |  |
| Symptom onset to groin puncture (minutes) | 1.00 | 0.99 – 1.00 | 0.077 |  |  |  |
| Baseline NIHSS | 0.95 | 0.90 – 0.99 | **0.043\*** | 1.03 | 0.96 – 1.12 | 0.402 |
| CBF | 0.99 | 0.98 – 1.01 | 0.878 |  |  |  |
| Site of Occlusion |  |  |  |  |  |  |
| ICA | Ref | Ref | Ref |  |  |  |
| M1 | 1.29 | 0.55 – 2.97 | 0.557 |  |  |  |
| TICI |  |  |  |  |  |  |
| 0-2A | Ref | Ref | Ref |  |  |  |
| 2B-3 | 3.40 | 1.25 – 9.23 | **0.016\*** | 4.11 | 1.10 – 15.31 | **0.035\*** |
| Discharge disposition |  |  |  |  |  |  |
| Home | Ref | Ref | Ref |  |  |  |
| Expired | 0.01 | 0.001 – 0.14 | **<0.001\*** | 0.03 | 0.002 – 0.44 | **0.010\*** |
| Hospice | 0.07 | 0.01 – 0.48 | **0.006\*** | 0.04 | 0.002 – 0.57 | **0.018\*** |
| Hospital or Rehab | 0.09 | 0.03 – 0.27 | **<0.001\*** | 0.13 | 0.03 – 0.45 | **0.002\*** |
| Atrial fibrillation | 0.57 | 0.28 – 1.18 | 0.133 |  |  |  |
| Hypertension | 0.43 | 0.20 – 0.91 | **0.026\*** | 0.61 | 0.23 – 1.61 | 0.320 |
| Type-2 diabetes | 0.72 | 0.30 – 1.76 | 0.476 |  |  |  |
| EF | 1.00 | 0.97 – 1.03 | 0.996 |  |  |  |
| Cervical Carotid involvement |  |  |  |  |  |  |
| No | Ref | Ref | Ref |  |  |  |
| Yes | 0.78 | 0.20 – 3.04 | 0.718 |  |  |  |
| ASPECTS |  |  |  |  |  |  |
| 2-5 | Ref | Ref | Ref |  |  |  |
| ≥6 | 0.59 | 0.13 – 2.79 | 0.507 |  |  |  |
| DAWN |  |  |  |  |  |  |
| True | Ref | Ref | Ref |  |  |  |
| False | 1.04 | 0.49 – 2.23 | 0.908 |  |  |  |
| Defuse |  |  |  |  |  |  |
| True | Ref | Ref | Ref |  |  |  |
| False | 1.45 | 0.67 – 3.16 | 0.348 |  |  |  |

\*Statistically significant at $p \leq 0.05$
Abbreviations: LOS= Length of Stay, ER= Emergency Room, TPA= Tissue Plasminogen Activator, SD=standard deviation, mRS=modified Rankin Score, IQR=interquartile range, NIHSS=national institute of health stroke scale, ICA=internal carotid artery, MCA=middle cerebral artery, CBF = Cerebral Blood Flow (ie, volume of ischemic tissue), mTICI = Modified Thrombolysis in Cerebral Infarction, ASPECTS=Alberta stroke program early computed tomography score, DEFUSE= DEFUSE 3 Trial, DAWN =DAWN Trial.

Notably, achieving higher levels of reperfusion (as indicated by a TICI score of 2B-3) emerged as a strong predictor of favorable outcomes, maintaining its significance in both univariate and multivariate analyses. Moreover, various discharge dispositions, particularly being discharged to home, were significantly associated with favorable outcomes in both univariate and multivariate analyses.

## Discussion

These data represent a single center, retrospective analysis that sought to identify clinical and imaging factors that could be utilized as prognostic indicators of favorable outcomes of endovascular stroke treatment in patients presenting with a proximal ICA/M1 MCA occlusion in the expanded thrombectomy time window. We acknowledge the limitations of our study-including our retrospective and single center design. Our single center model and low sample size was limiting in terms of statistical power for some analyses.

Our data mirrored findings of the DEFUSE-3 and DAWN study (Figure 2), demonstrating that later time from symptom onset to arterial groin puncture (>12 hours) was associated with a better odds ratio (OR 1.001; p-value 0.019) of an improved functional outcome (mRS 0-2) at 90 days compared with earlier arrival. This contrasts with other data suggesting that last known normal (LKN) to time to groin puncture was a significant prognostic indicator of a favorable outcome^12,13^. A potential explanation for this difference is that it may be a less critical factor in patients presenting in the expanded time window.

**Figure 2:**
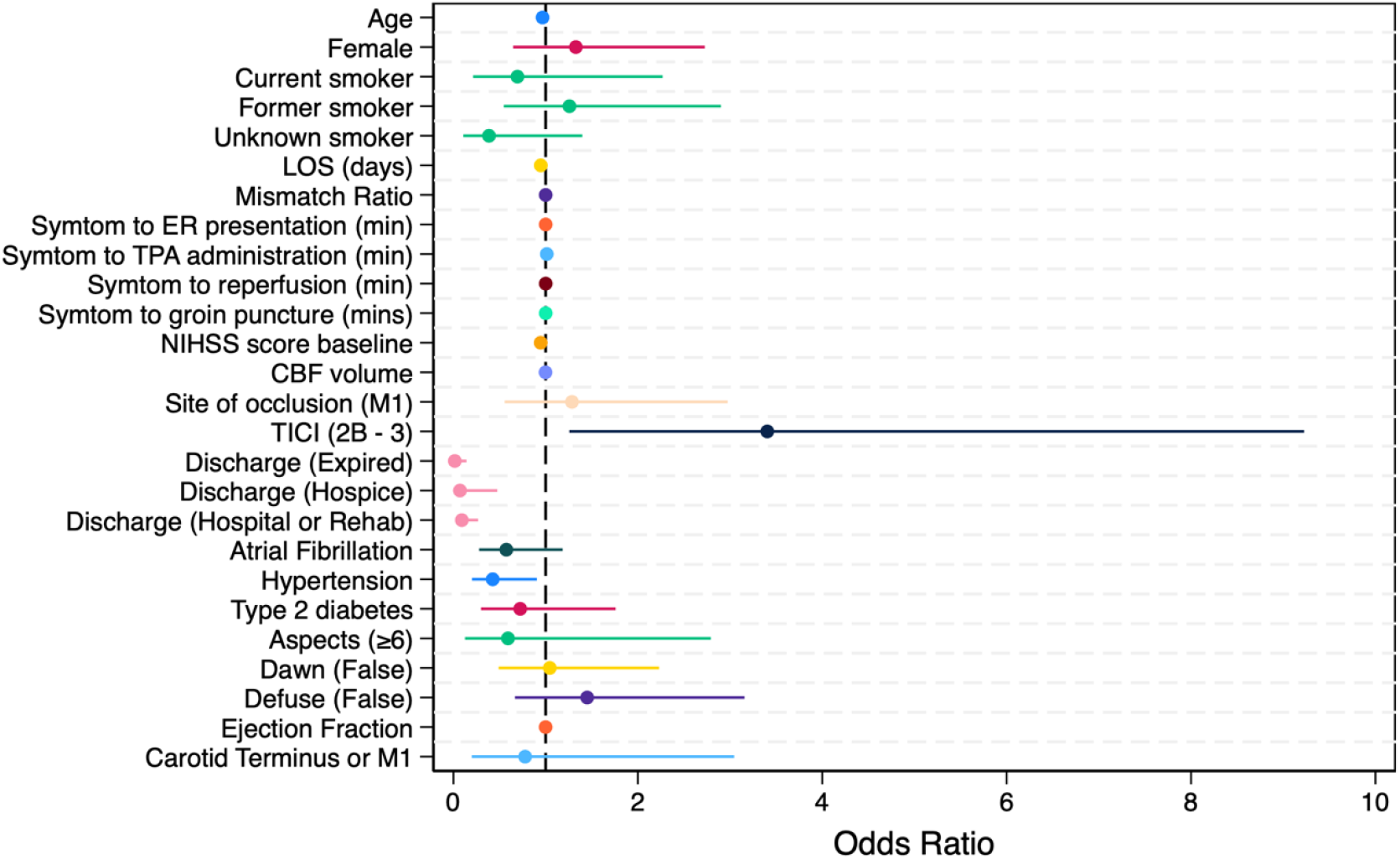
Forest plot of odds ratio of variables and their association with thrombectomy outcome.

Both DAWN and DEFUSE-3 demonstrated larger absolute benefits with later treatments compared with early window thrombectomy RCTs (e.g., HERMES, MR CLEAN, etc). This was later dubbed the “Late Window Paradox”^14^. Proposed explanations for this paradox include-slow growth of the ischemic core in some patients, presence of more robust collateral circulation, and the influence of lack of tPA administration in the medical control groups of the late window trials. It is possible that our population represented one with more robust collateral circulation.

This finding countered the popular mantra of “time is brain,” which implied that time to reperfusion was the sole variable important for LVO outcomes. Further, it demonstrated the collective heterogeneity of this patient population and cerebrovasculature. We hypothesize that presence of robust collateral circulation and delayed growth of the ischemic core have a significant influence on overall prognosis following endovascular stroke therapy. This implies that the degree of mismatch is the more important factor in patient selection for endovascular stroke treatment.

The AURORA study^15^, which was a pooled analysis of six RCTs (including DAWN and DEFUSE-3)-sought to answer the question regarding is there an advantage to selecting based off clinical mismatch (DAWN criteria), or target perfusion mismatch (DEFUSE-3 criteria). Importantly, it found a similar benefit in terms of functional independence (mRS 0-2) at 90 days regardless of which mismatch criteria was chosen. This finding was reflected in our data set that showed no statistical impact on outcome whether DAWN or DEFUSE-3 criteria were used.

Further research is investigating to what degree of mismatch is necessary for functional benefit to be achieved-but recent studies are demonstrating that benefit is possible beyond the mismatch criteria defined in the DAWN and DEFUSE-3 studies. Recently published RCTs, the SELECT-2^16^ and RESCUE-JAPAN LIMIT^17^ have demonstrated that patients with a low ASPECTs score (which is generally considered to correlate to the area of core infarct) have greater odds of achieving functional independence with MT compared with undergoing medical therapy alone. Both RCTs along with other observational studies and meta-analyses have already demonstrated the benefit of endovascular stroke treatment despite a low ASPECTs score (defined as ASPECTs 2-5)^18,7^. This is consistent with our data that showed no statistically significant association with ASPECTs score and a favorable endovascular stroke treatment outcome. Further, this also suggests that the degree of recanalization, as demonstrated by the post-procedural mTICI score is also the more critical indicator of prognosis which has also been shown in the literature.^19^

A significant negative association that our data reflected was that the presence of hypertension (defined as a prior to admission, SBP > 140) was less likely to lead to a favorable outcome (OR 0.32; p-value 0.009; 95% CI 0.12-0.75). This is also consistent with other multicenter data that demonstrates greater odds of a poor outcome with elevated pre-, peri-, and post-endovascular treatment blood pressure^20^. Possible explanations identified include an increased risk of symptomatic intracerebral hemorrhage (sICH) and reduced rates of successful recanalization.

Our data also demonstrated that the presence of symptomatic post-endovascular treatment ICH was more likely to be associated with a less favorable outcome and higher post-procedure mRS. This is consistent with other investigations that have demonstrated an association between hypertension and post-endovascular treatment sICH. However, the causality association was not investigated in our data, whether patients with pre-existing hypertension are more likely to have sICH is still an open question. This is an area of interest to be examined in future analyses.

The greater driving factor of this negative association may be more related to decreased odds of successful recanalization. A strong association between hypertension and increased vessel tortuosity is well established and could result in reduced ability to pass an angiocatheter to the site of occlusion and overall success of endovascular stroke treatment^21^. Another interesting hypothesis suggests that uncontrolled HTN may be inversely associated with the development of collateral flow. This would result in a greater hemodynamic force imparted on the area of occlusion and could potentially cause clot impaction and impaired ability for mechanical clot retrieval^22^.

Findings we expected demonstrated by our data include-younger age (p-value 0.007), shorter length of stay (p-value 0.003), and lower baseline NIHSS on presentation (p-value 0.021) had greater odds of a favorable outcome. Negative associations demonstrated included discharge to hospice was more likely to be associated with a poor functional outcome (mRS >0-2, p-value 0.001). This likely reflects that patients with a favorable functional outcome are more likely to be discharged home compared to hospice or rehab. Overall, these data add to a growing body of literature that works to refine criteria for patient selection for endovascular stroke treatment in the expanded thrombectomy time window.

## Data Availability

All data underlying the results are available as part of the article. No additional source data is required. Raw data used in analysis is available upon request.

## Acknowledgments, Sources of Funding, & Disclosures

**Acknowledgments**: The authors acknowledge Dr. Eric Adelman, for critically reviewing this manuscript.

## Sources of Funding

This work was supported by start-up funds, to B.F, from the University of Wisconsin School of Medicine and Public Health, and the department of Neurology University of Wisconsin, School of Medicine and Public Health, Madison, WI.

## Disclosures

The authors have nothing to disclose.

## Author Contributions

JC contributed to chart review, data acquisition, analysis, and authorship of the manuscript. QS contributed to chart review, data acquisition, analysis, and authorship of the manuscript. TR and TC contributed to data acquisition and drafting of methods. FH contributed data analysis and authorship of results. NB contributed to chart review, data acquisition, and drafting of methods. VN contributed to data analysis. VP contributed to study design, data acquisition, and clinical expertise. BAK contributed to study design, acquisition of clinical data, and clinical expertise. EA contributed to authorship and critical review of the manuscript. BF contributed to study conception, design, acquisition of clinical data, and clinical expertise. All authors of this study contributed to authorship and revision of the final manuscript.

## Author Contributions

JC contributed to chart review, data acquisition, analysis, and authorship of the manuscript. QS contributed to chart review, data acquisition, analysis, and authorship of the manuscript. TR and TC contributed to data acquisition and drafting of methods. FH contributed data analysis and authorship of results. NB contributed to chart review, data acquisition, and drafting of methods. EA critically reviewed and revised the manuscript and provided clinical expertise. VN contributed to data analysis. VP contributed to study design, data acquisition, and clinical expertise. BAK contributed to study design, acquisition of clinical data, and clinical expertise. BF contributed to study conception, design, acquisition of clinical data, and clinical expertise. All authors of this study contributed to authorship and revision of the final manuscript.

**Appendix**

## Notes

### Competing Interest Statement

The authors have declared no competing interest.

### Funding Statement

No external funding was received

### Author Declarations

Approved by University of Wisconsin SMPH IRB under Protocol ID: 2016-0418

